# Association of memory function with COVID-19 outcomes in adults aged 50 years and older: Analysis of three prospective cohorts

**DOI:** 10.1101/2025.01.24.25321065

**Authors:** Juanjuan Shi, Xin Shen, Yan Tian, Rui Lu, Jia Li, Xiaozhen Geng, Song Zhai, Xiaoli Jia, Fanpu Ji, Shuangsuo Dang, Wenjun Wang

## Abstract

**Background:** Patients with Alzheimer’s disease or dementia are at increased risk for COVID-19 hospitalization and mortality. However, no study has examined whether memory function is associated with COVID-19 outcomes in general older adults.

**Methods:** Data were obtained from SHARE (the Survey of Health, Ageing and Retirement in Europe), HRS (the Health and Retirement Study), and ELSA (the English Longitudinal Study of Ageing), three prospective and representative cohorts of non-institutionalized adults aged 50 years and older in 25 European countries plus Israel, the United States, and the United Kingdom, respectively. Memory function was measured with immediate and delayed 10-words recall tests. Associations of 10-words recall with COVID-19 hospitalization and mortality were assessed using logistic models adjusted for age, sex, race, body mass index, smoking, physical activity, household income, education level, and chronic conditions.

**Results:** A total of 4062 participants with COVID-19 infection from SHARE, 1349 from HRS, and 278 from ELSA were included in the analysis. 610 (15.0%) in SHARE, 142 (10.5%) in HRS, and 39 (14.0%) in ELSA were hospitalized, and 102 (2.5%) died of COVID-19 or related complications in SHARE. The adjusted odds ratios (aORs) for COVID-19 hospitalization were 1.15 (95% CI, 1.09-1.22) in SHARE, 1.07 (95% CI, 0.94-1.21) in HRS, and 1.34 (95% CI, 1.02-1.77) in ELSA, per word decrease in immediate 10-words recall. For delayed 10-words recall, the corresponding aORs were 1.11 (95% CI, 1.06-1.17), 1.12 (95% CI, 1.01-1.24), and 1.25 (95% CI, 1.01-1.55), respectively. The aORs for COVID-19 mortality were 1.07 (95% CI, 0.94-1.21) and 1.14 (95% CI, 1.01-1.28) per word decrease in immediate and delayed 10-words recall in SHARE, respectively. Results were relatively robust to missing data of covariates, exclusion of cases based on symptoms alone, or exclusion of cases with Alzheimer’s disease or dementia.

**Conclusions:** This study shows that low memory performance, as measured by 10-words recall, is independently associated with an increased risk of COVID-19 hospitalization and mortality in adults aged 50 years and older.

## Introduction

Coronavirus disease 2019 (COVID-19) has posed a significant global public health threat. As of March 2023, more than 760 million confirmed cases and over 6.8 million deaths have been reported worldwide.^1^ The clinical manifestations of COVID-19 range from asymptomatic cases to severe illness and death. Older adults are particularly at risk, with the majority of severe and fatal cases occurring in individuals aged 50 years and older.^2^ Hospitalization and mortality rates among this population are 3 to 15 times and 25 to 350 times higher, respectively, compared to younger individuals.^2^ Understanding the risk factors for severe and fatal COVID-19 outcomes in this vulnerable group is critical for identifying high-risk patients who may benefit from early intervention to prevent disease progression and adverse outcomes.

Established risk factors for severe COVID-19 include male sex, low socioeconomic status, poor physical fitness, and a range of chronic underlying conditions, such as cardiovascular disease, respiratory disease, cancer, obesity, diabetes, and dementia.^3–5^ However, despite these known factors, other potential contributors to severe COVID-19 outcomes remain to be fully understood.

One such potential risk factor is memory function, a key aspect of cognitive health that plays a central role in various other cognitive processes. Impaired memory has been linked to higher mortality in both the general population and in individuals with chronic illnesses, such as cancer and end-stage renal disease.^6–8^ While patients with Alzheimer’s disease or dementia have been shown to have an increased risk of COVID-19 hospitalization and mortality,^9–12^ it remains unclear whether poor memory performance in the general older adult population is similarly associated with adverse COVID-19 outcomes.

This study aims to fill this gap by examining the associations between pre-pandemic memory function and COVID-19 hospitalization and mortality in adults aged 50 years and older across three prospective cohorts. By investigating this hypothesized relationship, we seek to determine whether memory performance can serve as an additional risk factor for severe COVID-19 outcomes, which could have important implications for early risk identification and patient management.

## Methods

Data were obtained from SHARE (the Survey of Health, Ageing and Retirement in Europe),^13^ HRS (the Health and Retirement Study),^14^ and ELSA (the English Longitudinal Study of Ageing),^15^ three prospective and representative cohorts of non-institutionalized adults aged 50 years and older in 25 European countries plus Israel, the United States, and the United Kingdom, respectively. Details of the design and methods of the three cohorts can be found elsewhere.^13–15^ After the COVID-19 outbreak, participants in the three cohorts participated in surveys on COVID-19 infection and changes in life during the pandemic (two rounds from 2020 to 2021 in SHARE, two rounds from 2020 to 2022 in SHARE, and two rounds during 2020 in ELSA). SHARE, HRS, and ELSA were approved by the Ethics Council of the Max Planck Society, the Institutional Review Board of the University of Michigan, and the South Central Berkshire Research Ethics Committee, respectively.

We used data from wave 7 (2017) in SHARE, wave 14 (2018) in HRS, and wave 9 (2018/2019) in ELSA. These waves were considered baselines because they had the most recent memory tests before the COVID-19 outbreak. COVID-19 infection was defined if participants experienced COVID-19 symptoms, were tested positive for coronavirus, were told by a health care provider that they had the disease, attended an emergency department, or were hospitalized for COVID-19, or died of COVID-19 or related complications (Supplementary material, Appendix 1). Participants were excluded if they did not have 10-words recall tests at baseline, did not have COVID-19 infection, were younger than 50 years in 2020, or had missing values for covariates.

### COVID-19 hospitalization

In SHARE, participants were asked, “Who was hospitalized due to an infection from the Coronavirus?” Participants who reported being hospitalized were included in the COVID-19 hospitalization analysis. In HRS and ELSA, COVID-19 hospitalization was identified by the question “Were you admitted to the hospital because of the virus?” and “Have you had to stay in hospital for treatment due to coronavirus?”

### COVID-19 mortality

SHARE asked interviewers to verify a participant’s death by contacting a proxy respondent. If the participant was deceased, an end-of-life interview was undertaken to collect information related to the death. The proxy respondent was selected from the deceased participant’s immediate social network, including family/household members, neighbors, or other close acquaintances. Participants who died of COVID-19 or related complications were included in the COVID-19 mortality analysis. These participants also constituted the COVID-19 hospitalization sample, as they typically died in healthcare facilities. HRS and ELSA were not analyzed for COVID-19 mortality because their end-of-life data are not currently available.

### Memory function

Memory was assessed using the 10-words recall tests,^16^ which were conducted in a similar manner across the three cohorts. Participants were presented with a list of 10 words, one at a time. After the list was completed, participants were asked to recall and either type or speak as many words as they could remember, in any order (immediate 10-word recall). Approximately five minutes later, they were asked to recall the words again (delayed 10-word recall). The words used in the test were simple, everyday terms, such as “hotel,” “river,” “tree,” “skin,” “gold,” “market,” “paper,” “child,” “king,” and “book.” Scores on both the immediate and delayed recall tests ranged from 0 to 10, representing the number of words successfully remembered by the participants.

### Covariates

Potential confounders collected in all three cohorts were included in the analyses: age in 2020, sex, race (HRS and ELSA only), body mass index, smoking status, physical activity, household income, education, and underlying health conditions. These factors were previously reported to be related to COVID-19 outcomes.^3–5^ Body mass index was calculated as weight/height^2^, and height and weight were self-reported. Participants were asked how many days per week they did moderate and vigorous physical activity. If participants answered “rarely or never”, they were considered to be physical inactivity. Smoking status was divided into two categories: ever or current smoker and never smoker. In SHARE, educational attainment was coded based on the International Standard Classification of Education-97 (ISCED-97) and classified as low (no education or codes 1 and 2), medium (codes 3 and 4), and high (codes 5 and 6). In HRS and ELSA, low, medium, and high levels of education were less than high school, high school, and more than high school, respectively. Household income was categorized into country-specific quartiles. The following health conditions were asked whether participants had ever been diagnosed with or had at the time of the baseline interview: respiratory disease (such as emphysema or chronic bronchitis), cardiovascular disease (heart attack, congestive heart failure, or any other heart problem, and stroke or cerebral vascular disease), diabetes, cancer, and dementia or Alzheimer’s disease.

### Statistical analyses

Continuous variables were presented as the median and interquartile range (IQR). Categorical variables were presented as numbers and percentages. Differences in characteristics among the three cohorts were tested using the Kruskal-Wallis test or the chi-square test. Based on the results of these comparisons, we assessed the heterogeneity of the three cohorts and the suitability of pooling the results.

Four logistic regression models were fitted to test the associations of immediate and delayed 10-words recall with COVID-19 hospitalization and mortality. Model 0 was unadjusted. Model 1 adjusted for age, sex, and race (HRS and ELSA only). Model 2 additionally adjusted for body mass index, smoking status, physical activity, household income, and educational attainment. Finally, Model 3 further adjusted for underlying chronic diseases, including respiratory disease, cardiovascular disease, diabetes, cancer, and dementia or Alzheimer’s disease. Linear associations between continuous independent variables and COVID-19 hospitalization and mortality were tested using the Box-Tidwell test. As no evidence of deviation from linearity was found, the 10-words recall was treated as a continuous variable in the aforementioned logistic models, and odds ratios (ORs) per word decrease were calculated.

Three separate sensitivity analyses were performed. In the first sensitivity analysis, participants were excluded if the diagnosis of COVID-19 infection was based on symptoms alone. Thus, the included participants were those who were tested positive for coronavirus, were hospitalized due to COVID-19, or died of COVID-19 or related complications. In the second sensitivity analysis, participants were excluded if they had ever been diagnosed with or had dementia or Alzheimer’s disease at the time of the baseline interview. In the third sensitivity analysis, missing values of covariates were imputed (Supplementary material, Appendix 2). Due to the small sample size, the above sensitivity analyses were not performed for ELSA.

ORs were reported along with their 95% confidence intervals (95% CI). Statistically significant was defined as *p* <0.05 on both sides. StataSE 15 statistical software (StataCorp) was used for all analyses.

## Results

### Study population

A total of 4062 participants with COVID-19 infection from SHARE (25 European countries and Israel), 1349 from HRS (the United States), and 278 from ELSA (the United Kingdom) were included in the analysis. **Figure 1** shows the detailed participant selection process. Across all three cohorts, the participants included in the study were generally younger, had higher levels of education, and reported higher household incomes. Additionally, in the SHARE and HRS cohorts, participants were more likely to be female, overweight or obese, and less likely to engage in physical inactivity or have cardiovascular disease compared to those who were excluded (Supplementary material, Table S1-S3).

**Figure 1.**
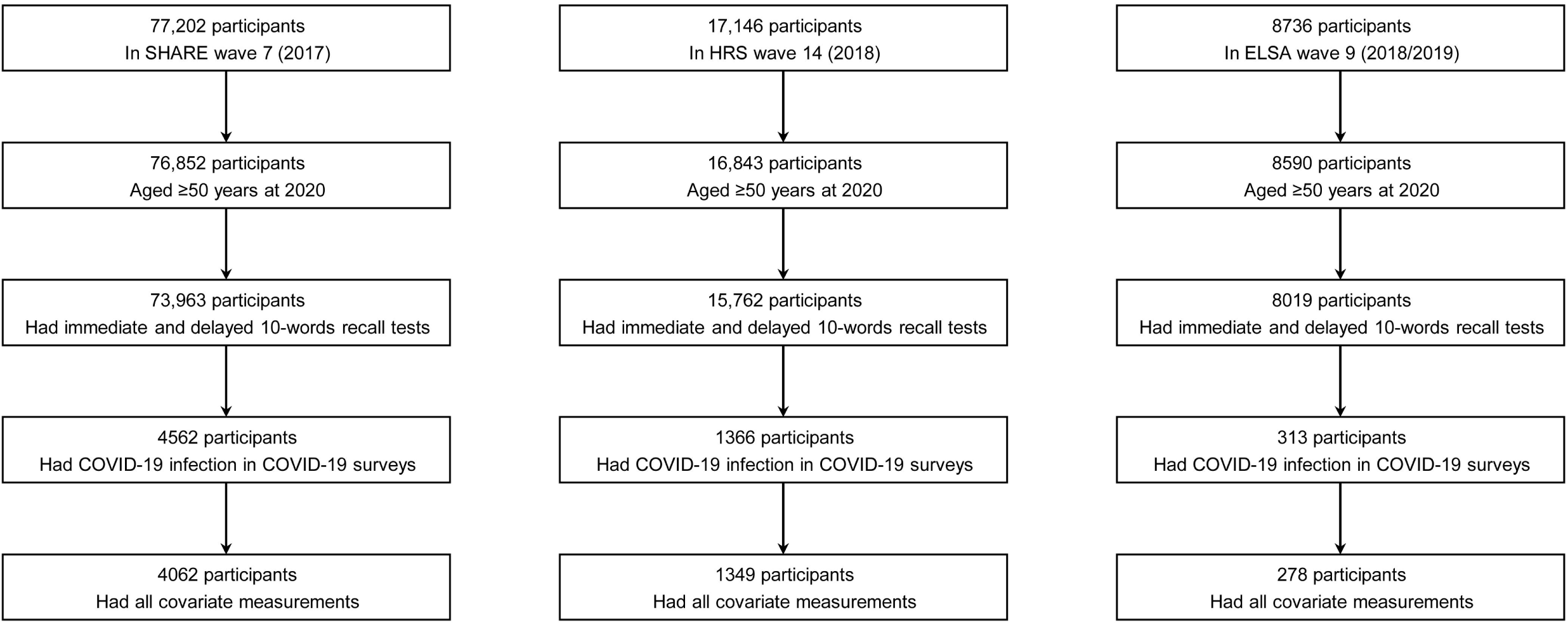
Determination of the study sample. COVID-19, coronavirus disease 2019; ELSA, the English Longitudinal Study of Ageing; HRS, the Health and Retirement Study; SHARE, the Survey of Health, Ageing and Retirement in Europe.

Median age of included participants was 67.0 (IQR: 62.0-74.0) years in SHARE, 64.0 (IQR: 59.0-71.0) years in HRS, and 66.0 (IQR: 57.0-73.0) years in ELSA. Men accounted for 40.9%, 42.0%, and 32.7% of the cohorts, respectively. The distributions of immediate and delayed 10-words recall test scores in the three cohorts are shown in **Figure 2**. The median number of words for immediate 10-words recall was 6 (IQR: 4-7) words in SHARE, 6 (IQR: 5-7) words in HRS, and 6.5 (IQR: 6-7) words in ELSA. For delayed 10-words recall, the corresponding median word counts were 4 (IQR: 3-6) words, 5 (IQR: 4-6) words, and 5 (IQR: 4-6) words, respectively. Most baseline characteristics differed among the three cohorts, as summarized in **Table 1**. Given the observed heterogeneity among the three cohorts, a pooled analysis was not performed.

**Figure 2.**
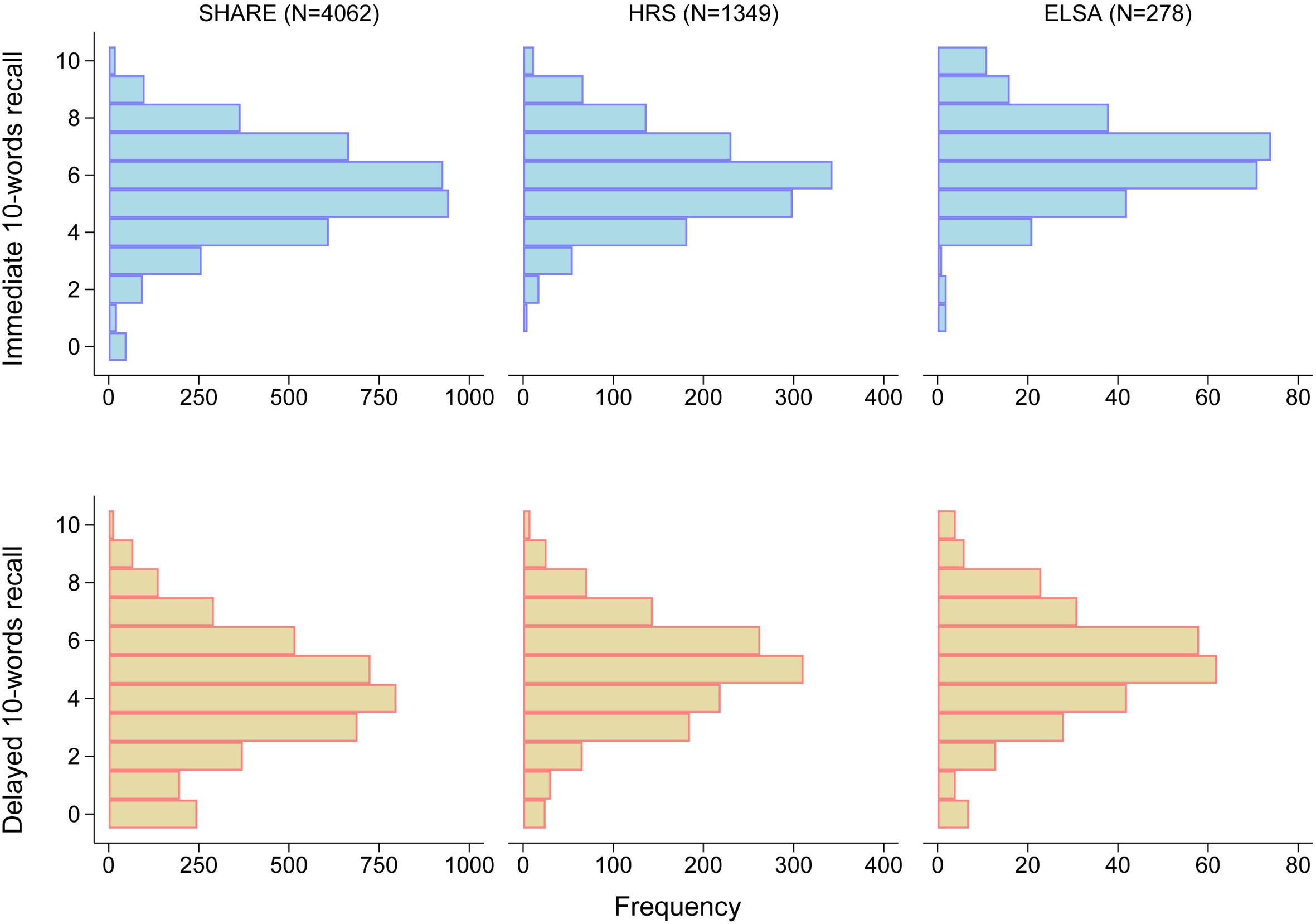
Distribution of scores on immediate and delayed 10-words recall tests. ELSA, the English Longitudinal Study of Ageing; HRS, the Health and Retirement Study; SHARE, the Survey of Health, Ageing and Retirement in Europe.

**Table 1.** Study Cohort Characteristics.

| Characteristic | SHARE<br>(n=4062) | HRS (n=1349) | ELSA (n=278) | P |
| --- | --- | --- | --- | --- |
| Age, median (IQR), y | 67.0 (62.0-74.0) | 64.0 (59.0-71.0) | 66.0 (57.0-73.0) | 0.0001 |
| Age category, y |  |  |  |  |
| 50-60 | 640 (15.8%) | 392 (29.1%) | 88 (31.7%) | <0.0001 |
| 60-70 | 1773 (43.7%) | 552 (40.9%) | 100 (36.0%) |  |
| 70-80 | 1139 (28.0%) | 259 (19.2%) | 58 (20.9%) |  |
| ≥80 | 510 (12.6%) | 146 (10.8%) | 32 (11.5%) |  |
| Sex |  |  |  |  |
| Male | 1660 (40.9%) | 566 (42.0%) | 102 (36.7%) | 0.264 |
| Female | 2402 (59.1%) | 783 (58.0%) | 176 (63.3%) |  |
| Race |  |  |  |  |
| White | NA | 903 (66.9%) | 265 (95.3%) | <0.0001 |
| Other | NA | 446 (33.1%) | 13 (4.7%) |  |
| Body mass index, kg/m<sup>2</sup> |  |  |  |  |
| Not overweight or obese: <25 | 1225 (30.2%) | 261 (19.4%) | 85 (30.6%) | <0.0001 |
| Overweight or obese: ≥25 | 2837 (69.8%) | 1088 (80.7%) | 193 (69.4%) |  |
| Educational level |  |  |  |  |
| Low | 1231 (30.3%) | 217 (16.1%) | 55 (20.0%) | <0.0001 |
| Medium | 1841 (45.3%) | 417 (30.9%) | 64 (23.0%) |  |
| High | 990 (24.4%) | 715 (53.0%) | 159 (57.2%) |  |
| Household income |  |  |  |  |
| Quartile 1 (lowest) | 795 (19.6%) | 282 (20.9%) | 60 (21.6%) | 0.050 |
| Quartile 2 | 991 (24.4%) | 289 (21.4%) | 56 (20.1%) |  |
| Quartile 3 | 1074 (26.4%) | 374 (27.7%) | 62 (22.3%) |  |
| Quartile 4 (highest) | 1202 (29.6%) | 404 (30.0%) | 100 (36.0%) |  |
| Ever or current smoker | 1708 (42.1%) | 691 (51.2%) | 159 (57.2%) | <0.0001 |
| Physical inactivity | 374 (9.2%) | 203 (15.1%) | 49 (17.6%) | <0.0001 |
| Cardiovascular disease | 548 (13.5%) | 335 (24.8%) | 22 (7.9%) | <0.0001 |
| Respiratory disease | 213 (5.2%) | 135 (10.0%) | 19 (6.8%) | <0.0001 |
| Diabetes | 510 (12.6%) | 388 (28.8%) | 34 (12.2%) | <0.0001 |
| Cancer | 175 (4.3%) | 187 (13.9%) | 34 (12.2%) | <0.0001 |
| Dementia or Alzheimer's disease | 42 (1.0%) | 24 (1.8%) | 1 (0.4%) | 0.039 |
| Immediate 10-words recall, median (IQR) words | 6 (4-7) | 6 (5-7) | 6.5 (6-7) | 0.0001 |
| Delayed 10-words recall, median (IQR) words | 4 (3-6) | 5 (4-6) | 5 (4-6) | 0.0001 |

Values are median (IQR) or n (%). Characteristics among the three cohorts were tested using the Kruskal-Wallis test or the chi-square test. ELSA, the English Longitudinal Study of Ageing; HRS, the Health and Retirement Study; SHARE, the Survey of Health, Ageing and Retirement in Europe.

### Associations of 10-word recall with COVID-19 hospitalization

Among participants with COVID-19 infection, 610 out of 4062 (15.0%) in SHARE, 142 out of 1349 (10.5%) in HRS, and 39 out of 278 (14.0%) in ELSA were hospitalized. Hospitalized participants recalled fewer words in the immediate 10-words recall test performed prior to the COVID-19 outbreak, compared with those who were not hospitalized, in each of the three cohorts ([median (IQR)]: 5 (4-6) words vs. 6 (5-7) words, in SHARE; 5 (4-7) words vs. 6 (5-7) words, in HRS; 6 (5-6) words vs. 7 (6-8) words, in ELSA; Supplementary material, Figure S1). The results were similar for the delayed 10-words recall test ([median (IQR)]: 3 (2-5) words vs. 4 (3-6) words, in SHARE; 4 (3-6) words vs. 5 (4-6) words, in HRS; 4 (3-5) words vs. 5 (4-7) words, in ELSA; Supplementary material, Figure S1).

As shown in **Table 2**, in Model 0, the crude ORs of hospitalization per word decrease in immediate 10-words recall were 1.31 (95% CI, 1.24-1.37) in SHARE, 1.23 (95% CI, 1.11-1.38) in HRS, and 1.54 (95% CI, 1.22-1.94) in ELSA. For delayed 10-words recall, the corresponding crude ORs per word decrease were 1.24 (95% CI, 1.19-1.30), 1.25 (95% CI, 1.14-1.37), and 1.43 (95% CI, 1.19-1.71), respectively. These ORs were dwarfed in each cohort, after adjustment for age, sex, and race in Model 1; after further adjustment, the magnitude of the ORs were slightly attenuated in Model 2 and Model 3. In the fully adjusted model (Model 3), the ORs for hospitalization were 1.15 (95% CI, 1.09-1.22) in SHARE, 1.07 (95% CI, 0.94-1.21) in HRS, and 1.34 (95% CI, 1.02-1.77) in ELSA, per word decrease in immediate 10-words recall (Supplementary material, Figure S2). For delayed 10-words recall in Model 3, the corresponding adjusted ORs per word decrease were 1.11 (95% CI, 1.06-1.17), 1.12 (95% CI, 1.01-1.24), and 1.25 (95% CI, 1.01-1.55), respectively (Supplementary material, Figure S3). Other factors associated with COVID-19 hospitalization were older age (SHARE and HRS), male sex (SHARE and HRS), non-white race (HRS), being overweight or obese (SHARE and HRS), physical inactivity (SHARE and HRS), lower levels of income (HRS and ELSA), lower levels of education (SHARE), diabetes (SHARE), and dementia or Alzheimer’s disease (HRS) (Supplementary material, Figure S2-S3).

**Table 2.**
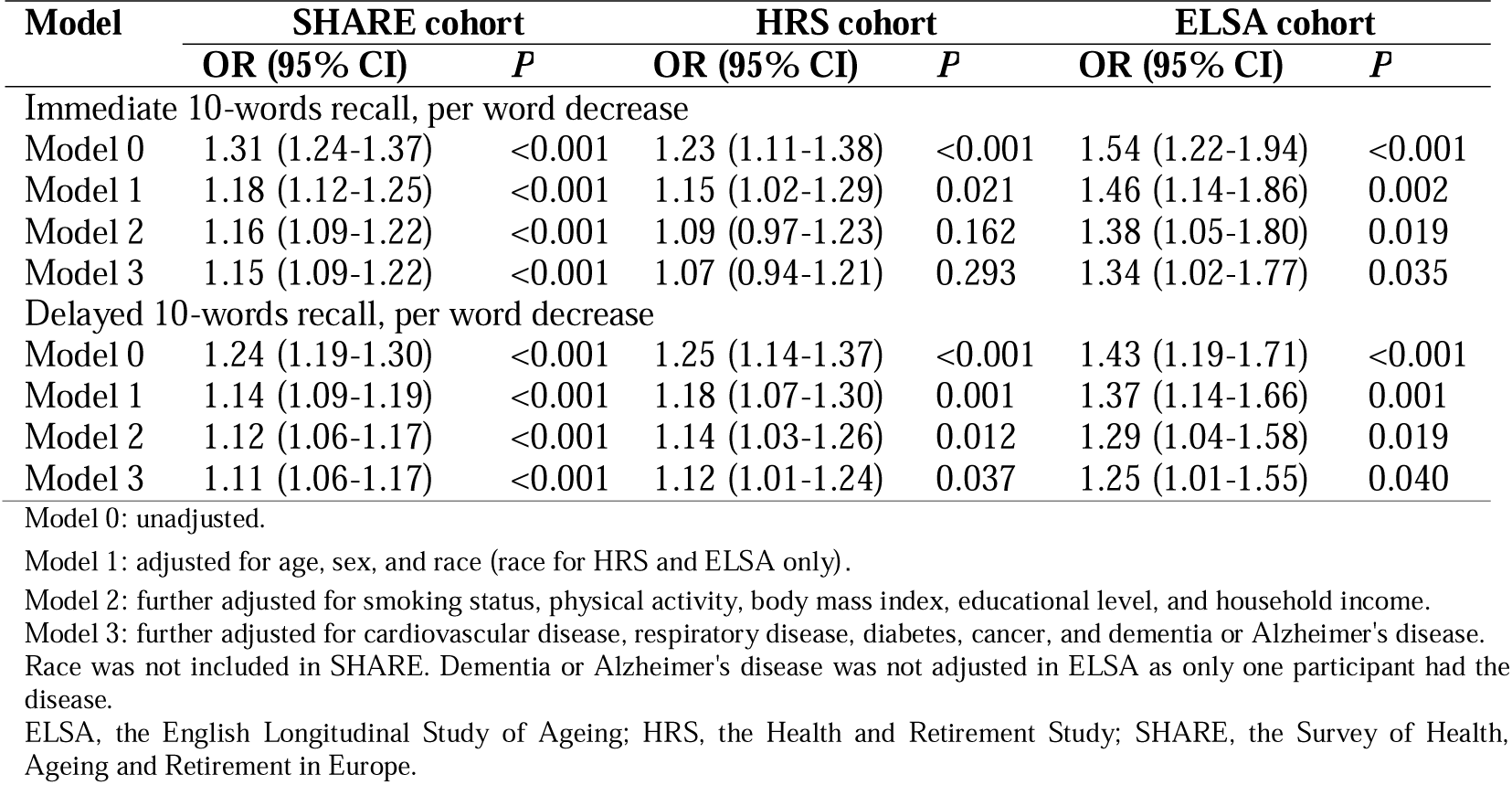
Associations of 10-words recall and COVID-19 hospitalization in three cohorts.

### Associations of 10-word recall with COVID-19 mortality

Only SHARE had end-of-life data to allow analysis of the COVID-19 mortality. Of the 4062 participants with COVID-19 infection in SHARE, 102 (2.5%) died of COVID-19 or related complications. Participants who died of COVID-19 or related complications recalled fewer words in both immediate and delayed recall tests performed before the COVID-19 outbreak compared with those who did not (immediate 10-words recall [median (IQR)]: 4.5 (3-6) words vs. 6 (5-7) words; delayed 10-words recall [median (IQR)]: 3 (1-4) words vs. 4 (3-6) words; Supplementary material, Figure S4).

As shown in **Table 3**, the crude ORs for COVID-19 mortality per word decrease were 1.40 (95% CI, 1.26-1.55) for immediate 10-words recall and 1.41 (95% CI, 1.28-1.56) for delayed 10-words recall. After full adjustment, the association with COVID-19 mortality was still statistically significant for delayed 10-words recall (OR, 1.14; 95% CI, 1.01-1.28; Supplementary material, Figure S5) but not for immediate 10-words recall (OR, 1.06; 95% CI, 0.93-1.20; Supplementary material, Figure S6). Other factors associated with COVID-19 mortality in SHARE were older age, male sex, physical inactivity, and diabetes (Supplementary material, Figure S5-S6).

**Table 3.** Associations of 10-words recall and COVID-19 mortality in the SHARE cohort.

| **Model** | **OR (95% CI)** | ***P*** |
| --- | --- | --- |
| Immediate 10-words recall, per word decrease |  |  |
| Model 0 | 1.40 (1.26-1.55) | <0.001 |
| Model 1 | 1.12 (1.00-1.26) | 0.055 |
| Model 2 | 1.07 (0.94-1.21) | 0.305 |
| Model 3 | 1.06 (0.93-1.20) | 0.409 |
| Delayed 10-words recall, per word decrease |  |  |
| Model 0 | 1.41 (1.28-1.56) | <0.001 |
| Model 1 | 1.19 (1.06-1.32) | 0.002 |
| Model 2 | 1.15 (1.02-1.29) | 0.020 |
| Model 3 | 1.14 (1.01-1.28) | 0.031 |

Model 0: unadjusted.
Model 1: adjusted for age and sex.
Model 2: further adjusted for smoking status, physical activity, body mass index, educational level, and household income.
Model 3: further adjusted for cardiovascular disease, respiratory disease, diabetes, cancer, and dementia or Alzheimer's disease. ELSA, the English Longitudinal Study of Ageing.

### Sensitivity analyses

After imputation for missing data of covariates, the association of delayed 10-words recall with COVID-19 mortality was no longer statistically significant in SHARE (OR, 1.09; 95% CI, 0.98-1.22). In other sensitivity analyses, there was little change in the magnitude of the associations with either immediate or delayed 10-word recall in both SHARE and HRS (Supplementary material, Table S4-S8).

## Discussion

The main finding of this study is that low pre-pandemic memory performance was associated with an increased risk of COVID-19-related hospitalization among individuals aged 50 years and older, based on data from three longitudinal cohorts across 25 European countries and Israel, the United States, and the United Kingdom. In addition, low pre-pandemic memory performance was linked to a higher risk of COVID-19 mortality in individuals from 25 European countries and Israel. These associations remained robust even after adjusting for established risk factors for severe COVID-19 outcomes, including older age, male sex, non-white race, low socioeconomic status, physical inactivity, and pre-existing chronic conditions such as cardiovascular disease, respiratory disease, cancer, diabetes, and obesity. Notably, the strongest association with hospitalization was observed in the delayed 10-word recall test, with consistent results across all three cohorts. In contrast, the immediate 10-word recall appeared less relevant, as its association with hospitalization was only observed in the SHARE and ELSA cohorts, but not in the HRS cohort.

To our best knowledge, rare study examined the association between memory function and COVID-19 outcomes in general older adults. Previous studies have focused primarily on patients with Alzheimer’s disease or dementia, which has been shown to be an independent risk factor for COVID-19 hospitalization and mortality.^9–12^ In this study, we also found that individuals with Alzheimer’s disease or dementia tend to be hospitalized or die when infected with COVID-19, especially for hospitalization in the HRS cohort. However, in our study, the samples were from community-dwelling populations. Alzheimer’s disease or dementia accounted for only a very small proportion of the study samples (<2%); the vast majority were older adults without dementia. Furthermore, we performed adjusted analyses and sensitivity analyses excluding Alzheimer’s disease or dementia. Therefore, we found the associations not only in general older adults, but also in the non-dementia older adults.

Recommendations for the management of COVID-19 are highly dependent on the risk of a patient progressing from a milder state to the next more severe state.^17^ The identification of risk factors for severe COVID-19 is of great importance to clinicians and to public health strategies for the management of this disease. In this study, we identified low memory performance, measured as 10-words recall test, as a potential risk factor of such. The test can be easily performed, and the respondent burden is low. Moreover, the test has been demonstrated good validity and reliability and now has been widely adopted, particularly in large surveys of aging populations.^13–15^ Its utility for COVID-19 prognosis and treatment guidance, especially in combination with established risk factors, is worthy of evaluation in both community and clinical settings.

Behavioral pathways and socioeconomic status are thought to mediate, in part, the association between memory function and clinical outcomes. For example, individuals with low memory performance are less likely to engage in healthy behaviors (e.g., healthy eating, physical activity, reduced alcohol consumption, and smoking cessation) and to adhere to their medication regimens.^18,19^ One study found that older adults with low memory performance were less likely to adhere to COVID-19 protective behaviors.^20^ The association between low socioeconomic status and predementia and dementia syndromes is supported by the majority of studies.^19^ In the current study, the ORs for COVID-19 hospitalization and mortality were attenuated after adjustment for smoking status, physical activity, body mass index, educational level, and household income (model 2), suggesting that health behaviors and socioeconomic status contribute to some extent to their associations.

Common factors, such as comorbidities, gut microbes, environmental factors, and inflammatory cytokines, can cause both low memory performance and severe disease in COVID-19. More than 100 diseases cause memory impairment.^21^ In this study, the ORs for COVID-19 hospitalization and mortality were attenuated after adjustment for underlying diseases, including cardiovascular disease, respiratory disease, diabetes, cancer, and obesity, suggesting that comorbidities contribute in part to the observed associations. Gut microbiota may alter brain integrity and cognitive function.^22^ One study showed that COVID-19 patients with lower gut microbiota diversity were more likely to require ICU admission.^23^ Adverse environments, such as air pollution, are associated with impaired cognition. A meta-analysis showed that 127 out of 139 studies reported an association between air pollution and unfavorable COVID-19 outcomes.^24^ Increased levels of inflammation, such as interleukin-1β, interleukin-6, and C-reactive protein, which can potentially cause or aggravate biological ageing and cellular ageing, are involved in memory loss and other cognitive functions.^25^ These inflammatory factors mediate COVID-19 progression and are well-defined markers of severe COVID-19.^26^

In addition to low memory performance, this study identified several factors associated with COVID-19 hospitalization. These include older age, male sex, non-white race, overweight or obesity, lower levels of education and household income, physical inactivity, and underlying conditions such as diabetes and dementia or Alzheimer’s disease. Furthermore, older age, male sex, physical inactivity, and diabetes were found to be associated with COVID-19 mortality. These are well-established risk factors for COVID-19 severity and death. However, other commonly recognized risk factors, such as smoking, cardiovascular disease, and chronic respiratory disease, were not found to be significant in this study. It is important to note that prior research has predominantly focused on individuals at the highest risk, often those already hospitalized. As a result, the findings of previous studies may not be generalizable to the broader population, as is the case in the present study.

The main strength of the study is the longitudinal design of SHARE, HRS, and ELSA, and their good representation of European, American, and British populations aged 50 years and older. The association of low memory performance with COVID-19 hospitalization was robust in our study. First, the association was observed in each of the three cohorts, even though there was great heterogeneity among the three cohorts. Second, the association was still observed after full adjustment for demographics, socioeconomic status, lifestyle, and underlying diseases. Third, the ORs for COVID-19 hospitalization remained largely unchanged in the three sensitivity analyses.

This study has several limitations. First, information on COVID-19 infection, hospitalization, mortality, and covariates was collected via questionnaires. This may introduce bias at various stages of questionnaire design and administration. Direct measurements or data linked to medical records and mortality registries would likely provide more accurate estimates of the associations. Second, although adjustments were made for a range of demographic, lifestyle, socioeconomic, and clinical factors, the potential influence of unmeasured variables (e.g., unadjusted comorbidities, gut microbiota, environmental factors, and inflammatory cytokines) cannot be entirely ruled out. Factors that were not available across all three datasets or that were measured using inconsistent methods, such as loneliness and social isolation,^27^ were also not adjusted. Advance care planning decisions (e.g., “do not resuscitate” or “do not intubate”)—factors known to affect severity and death—were not collected in the SHARE cohort, and their inclusion might have weakened or negated the observed associations. Third, the lack of end-of-life data from HRS and ELSA prevented the analysis of COVID-19 mortality in these two cohorts. Although SHARE provided mortality data, the number of COVID-19-related deaths was relatively small, and the results were sensitive to imputations for missing covariates. While SHARE had the largest sample size for hospitalization analysis, ELSA included only 39 hospitalized individuals. Older participants recalled fewer words across all three cohorts (data not shown), but due to the limited sample size in each age group, age-stratified analyses were not conducted. The imbalance between hospitalized and non-hospitalized participants may also have biased the results. Therefore, cohorts with larger sample sizes are needed to validate our findings. Fourth, there was a lack of data on COVID-19 vaccination status and viral strains, both of which are critical determinants of disease severity.^28^ The impact of these factors on the observed associations remains unclear, and updated data will be necessary as vaccination coverage increases and the virus continues to evolve.

### Conclusions

Our study shows that low memory performance, as measured by 10-words recall test, is independently associated with an increased risk of COVID-19 hospitalization and mortality in adults aged 50 years and older. Further studies are warranted to validate these associations in recent omicron waves of the COVID-19 pandemic and in clinical settings, and to elucidate the underlying mechanisms in order to improve the management of COVID-19.

## List of abbreviations

CI: confidence intervals
COVID-19: Coronavirus disease 2019
ELSA: the English Longitudinal Study of Ageing
HRS: the Health and Retirement Study
IQR: interquartile range
ISCED-97: International Standard Classification of Education-97
OR: odds ratio
SHARE: the Survey of Health, Ageing and Retirement in Europe

## Declarations

### Ethics approval and consent to participate

SHARE, HRS, and ELSA were approved by the Ethics Council of the Max Planck Society, the Institutional Review Board of the University of Michigan, and the South Central Berkshire Research Ethics Committee, respectively. Written informed consent was obtained from all the participants in SHARE, HRS, and ELSA.

### Consent for publication

Not applicable.

### Availability of data and materials

The datasets are freely available for download at the study websites (SHARE: http://www.shareproject.org/data-access.html; HRS: https://hrs.isr.umich.edu/; ELSA: https://www.elsa-project.ac.uk/).

### Authors’ contributions

JS, XS, YT, and WW conceived of and designed the study. WW and JS acquired the data and conducted the statistical analyses. XS and TY verified the data. All authors interpreted the data. JS, XS, and YT drafted the manuscript. All authors reviewed and contributed revisions to the final version of the paper. All authors approved the final version of the paper.

## Data Availability

The datasets are freely available for download at the study websites (SHARE: http://www.shareproject.org/data-access.html; HRS: https://hrs.isr.umich.edu/; ELSA: https://www.elsa-project.ac.uk/). Written informed consent was obtained from all the participants in SHARE, HRS, and ELSA.

## Acknowledgements

This paper uses data from SHARE. The SHARE data collection has been funded by the European Commission, DG RTD through FP5 (QLK6-CT-2001-00360), FP6 (SHARE-I3: RII-CT-2006-062193, COMPARE: CIT5-CT-2005-028857, SHARELIFE: CIT4-CT-2006-028812), FP7 (SHARE-PREP: no. 211909, SHARE-LEAP: no. 227822, SHARE M4: no. 261982, DASISH: no. 283646) and Horizon 2020 (SHARE-DEV3: no. 676536, SHARE-COHESION: no. 870628, SERISS: no. 654221, SSHOC: no. 823782, SHARE-COVID19: no. 101015924) and by DG Employment, Social Affairs & Inclusion through VS 2015/0195, VS 2016/0135, VS 2018/0285, VS 2019/0332, and VS 2020/0313. Additional funding from the German Ministry of Education and Research, the Max Planck Society for the Advancement of Science, the U.S. National Institute on Aging (U01_AG09740-13S2, P01_AG005842, P01_AG08291, P30_AG12815, R21_AG025169, Y1-AG-4553-01, IAG_BSR06-11, OGHA_04-064, HHSN271201300071C, RAG052527A) and from various national funding sources is gratefully acknowledged (see www.share-project.org). This paper uses data from HRS, sponsored by the National Institute on Aging (National Institutes of Health, Bethesda, MD; grant number NIA U01AG009740) and conducted by the University of Michigan (Ann Arbor, MI). This paper uses data from ELSA Wave 9, and COVID-19 wave 1 and 2. ELSA is funded by the National Institute on Aging (R01AG017644), and by UK Government Departments coordinated by the National Institute for Health and Care Research (NIHR).

