## Supplementary material for "Association of memory function with COVID-19 outcomes in adults aged 50 years and older: Analysis of three prospective cohorts"

Juanjuan Shi^1^; Xin Shen^2^; Yan Tian^1^; Rui Lu^1^; Jia Li^1^; Xiaozhen Geng^1^; Song Zhai^1^; Xiaoli Jia^1^; Fanpu Ji^1^; Shuangsuo Dang^1^; Wenjun Wang^1^

^1^ Department of Infectious Diseases, Second Affiliated Hospital of Xi’an Jiaotong University, 157 Xiwu Road, Xi’an 710004, China

^2^ Department of Medical Affairs, Shaanxi Provincial people’s Hospital, 256 West Youyi Road, Xi’an 710068, China

**Appendix 1. Definition of COVID-19 infection in SHARE, HRS, and ELSA**

In SHARE, COVID-19 infection was considered if a participant answered "Respondent" to any of the following questions, or the cause of death was COVID-19 or related complications in the end-of-life interview for a deceased participant:

CAC003 (Round 1): "Since the outbreak of Corona, who experienced symptoms that you would attribute to the Covid illness, e.g., cough, fever, or difficulty breathing?"

CAC005 (Round 1): "Who was tested positive for the Corona virus?"

CAC011 (Round 1): "Who was hospitalized due to an infection from the Corona virus?"

CAC103 (Round 2): "Since your last interview, who experienced symptoms that you would attribute to the Covid illness, e.g., cough, fever, difficulty breathing, or loss of sense of taste or smell?"

CAC105 (Round 2): "Who was tested positive for the Corona virus?"

CAC111 (Round 2): "Who was hospitalized due to an infection from the Corona virus?"

In HRS, COVID-19 infection was considered if a participant answered "Yes" to any of the following questions:

W551 (Round 1): "Have you had or do you now have COVID-19, the disease caused by the novel coronavirus?"

W553 (Round 1): "Did the test indicate that you had the virus?"

W557 (Round 1): "Did a doctor or other health care provider tell you that you have the disease?"

W558 (Round 1): "Did you have to go to an emergency room because of the virus?"

W559 (Round 1): "Were you admitted to a hospital because of the virus?"

A1_21S (Round 2): "Have you had, or do you now have, COVID-19? "

A2_21S (Round 2): "Did a doctor or other health care provider tell you that you had COVID-19?"

A11_21S (Round 2): "Did the test (or any of the tests if you had more than one), indicate that you had COVID-19?"

B1_21S (Round 2): "Did you have to go to the emergency room because of the virus?"

B2_21S (Round 2): "Were you admitted to the hospital because of the virus?"

In ELSA, COVID-19 infection was considered if a participant's response met any of the following:

CvSelfWhy1 (Round 1): "I have or had symptoms of coronavirus." to the question "Why were you self-isolating?"

CvTestB (Round 1): "It was positive." to the question "What was the result of your coronavirus test?)"

CvHosp (Round 1): "Yes" to the question “Have you had to stay in hospital for treatment due to coronavirus?"

CvTestB (Round 2): "Positive" to the question "What was the result of your coronavirus test?"

CvHosp (Round 2): "Yes" to the question "Since we last interviewed you have you had to stay in hospital for treatment due to coronavirus?"

CvLongCovid (Round 2): "Yes" or "No" to the question "Have you been told by a doctor that you have any long-standing illness or disability caused by coronavirus?"

**Appendix 2. Imputation methods for missing covariates in SHARE and HRS**

In total, 500 participants (11.0%) in the SHARE cohort and 17 (1.2%) in the HRS cohort were excluded from the main analyses due to missing values of covariates. In the sensitivity analyses, missing values for education level were imputed using their mean values. Then, a multiple imputation method using chained equations was used to impute missing values of other covariates. In SHARE, these imputed covariates were physical activity, smoking status, body mass index (overweight/obese or not), cardiovascular disease, respiratory disease, diabetes, cancer, and Alzheimer's disease or dementia, and in HRS were smoking status and body mass index (overweight/obese or not). In the chained equations, the logistic method was used as the univariate method, and age and gender were input as independent variables.

**Figure S1. Immediate and delayed 10-words recall tests by COVID-19 hospitalization in SHARE, HRS, and ELSA.**

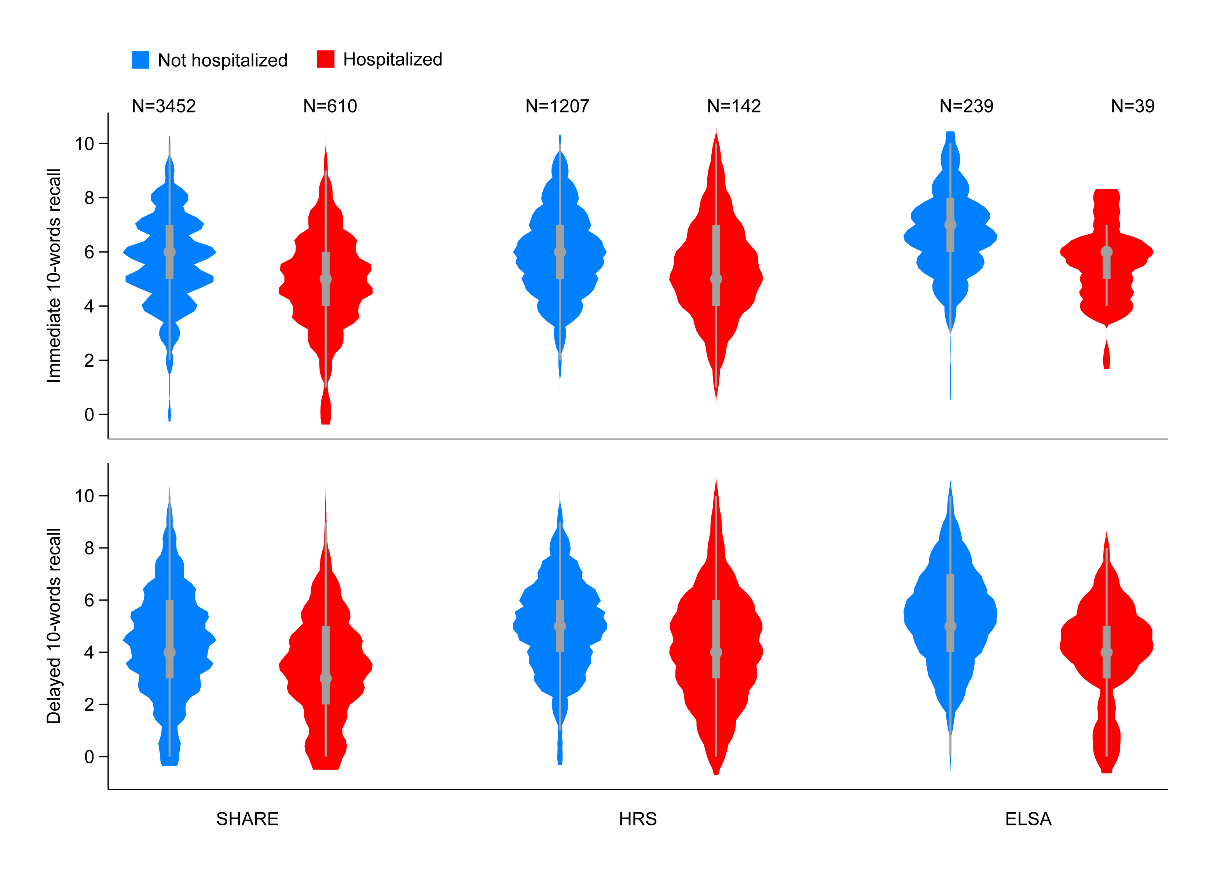

ELSA, the English Longitudinal Study of Ageing; HRS, the Health and Retirement Study; SHARE, the Survey of Health, Ageing and Retirement in Europe.

**Figure S2. Logistic regression for the association of immediate 10-words recall with COVID-19 hospitalization in SHARE, HRS, and ELSA.**

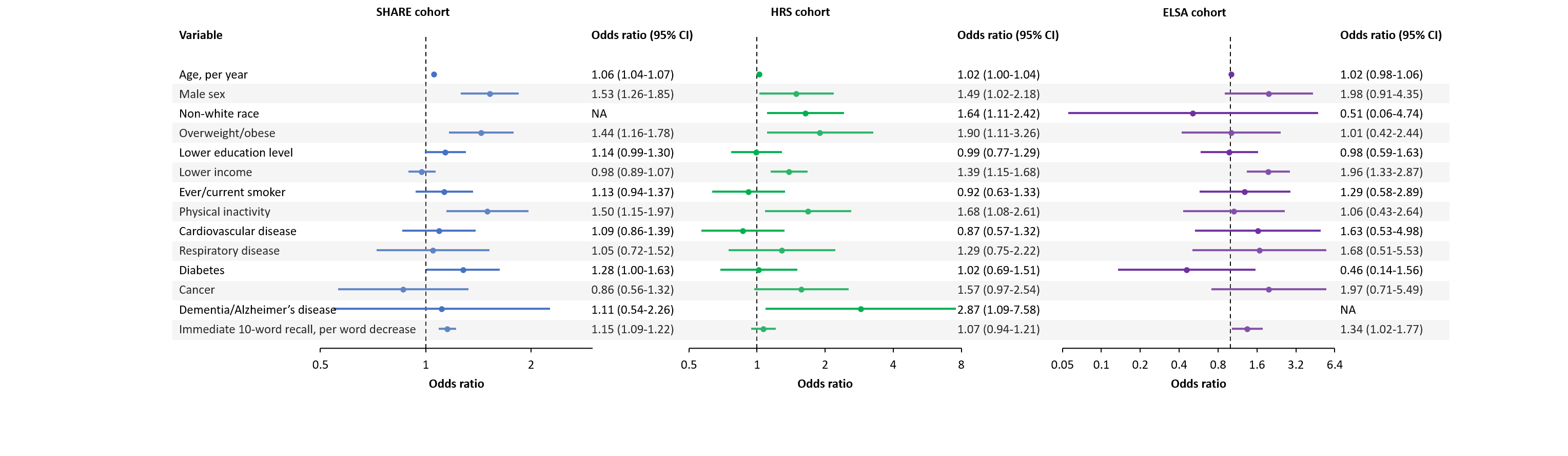

ELSA, the English Longitudinal Study of Ageing; HRS, the Health and Retirement Study; SHARE, the Survey of Health, Ageing and Retirement in Europe.

**Figure S3. Logistic regression for the association of delayed 10-words recall with COVID-19 hospitalization in SHARE, HRS, and ELSA.**

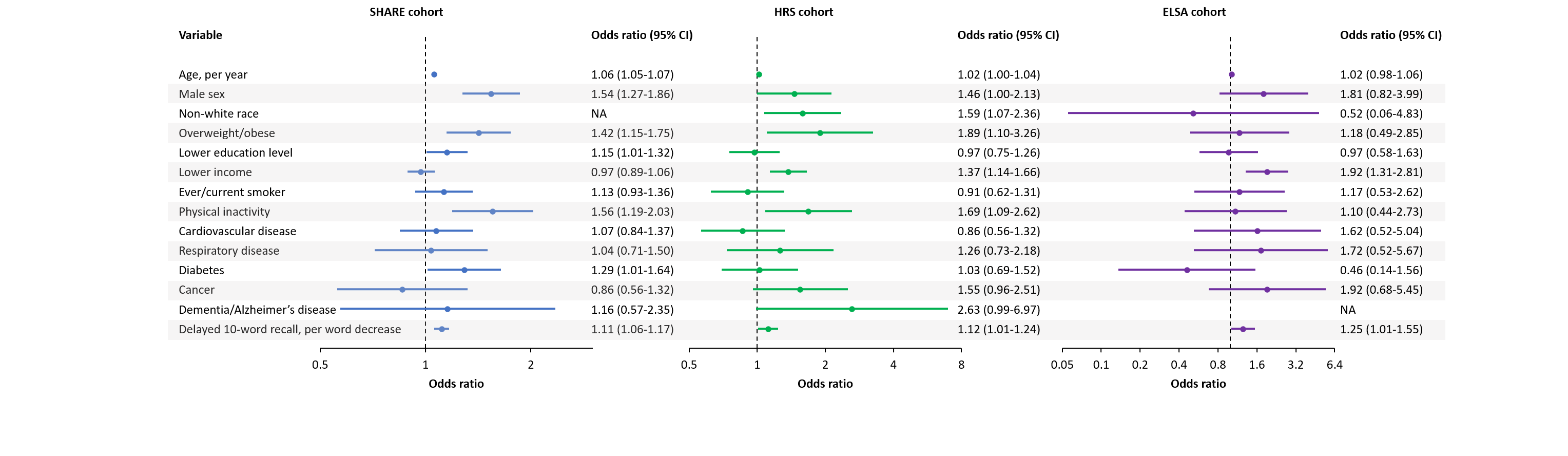
ELSA, the English Longitudinal Study of Ageing; HRS, the Health and Retirement Study; SHARE, the Survey of Health, Ageing and Retirement in Europe.

**Figure S4. Immediate and delayed 10-words recall tests by COVID-19 mortality in SHARE.**

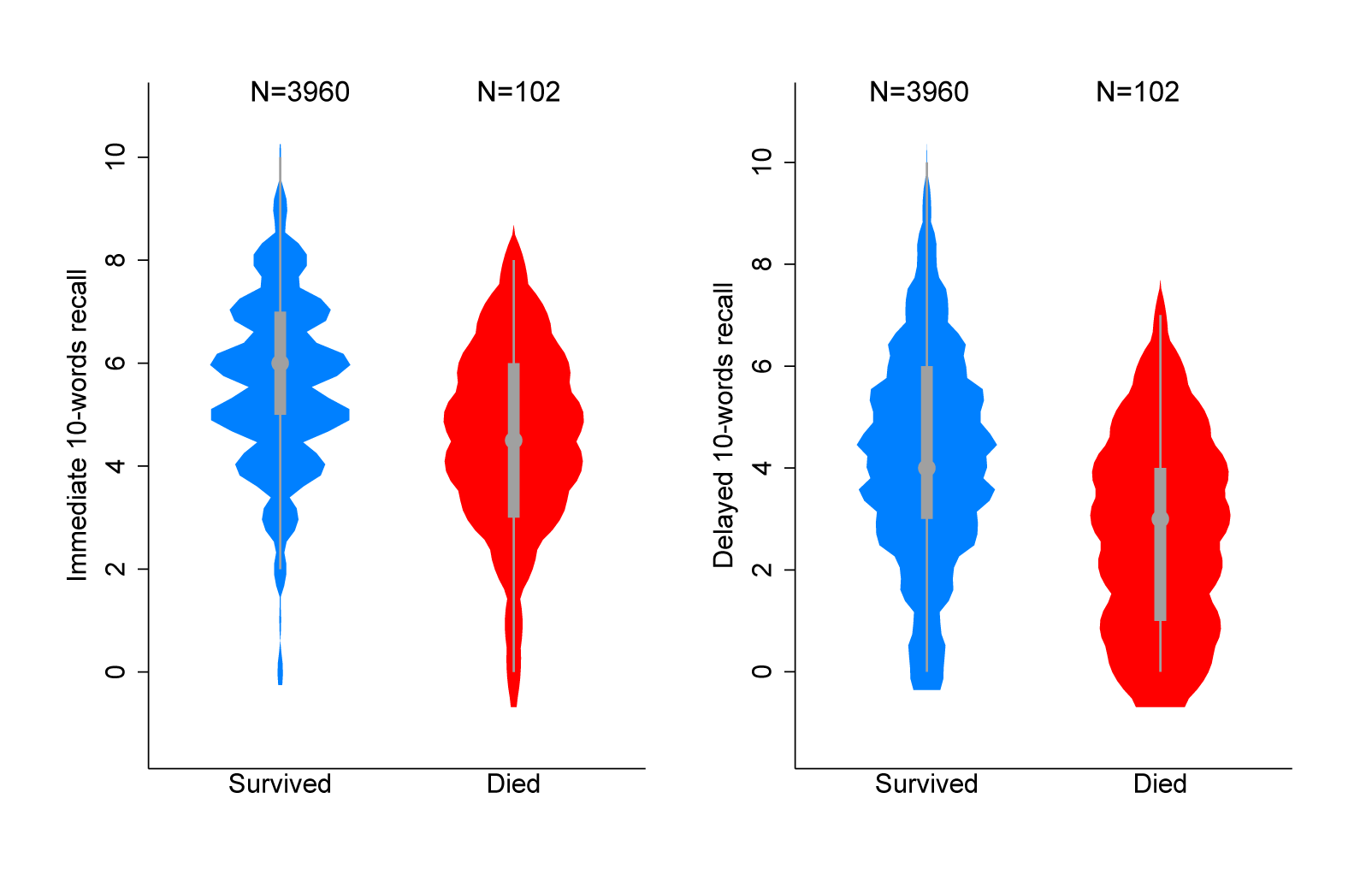

SHARE, the Survey of Health, Ageing and Retirement in Europe.

**Figure S5. Logistic regression for the association of delayed 10-words recall with COVID-19 mortality in SHARE.**

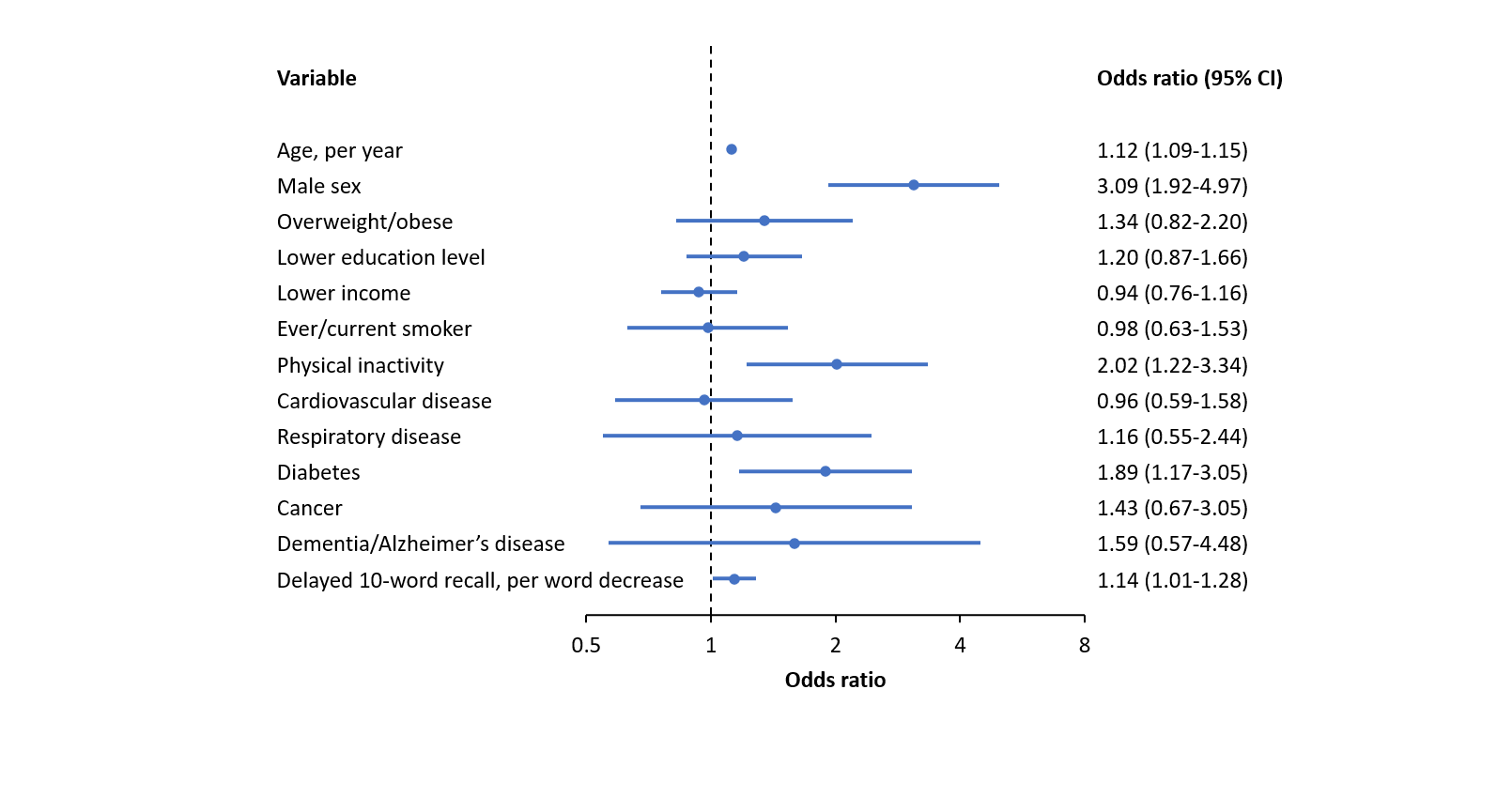

SHARE, the Survey of Health, Ageing and Retirement in Europe.

**Figure S6. Logistic regression for the association of immediate 10-words recall with COVID-19 mortality in SHARE.**

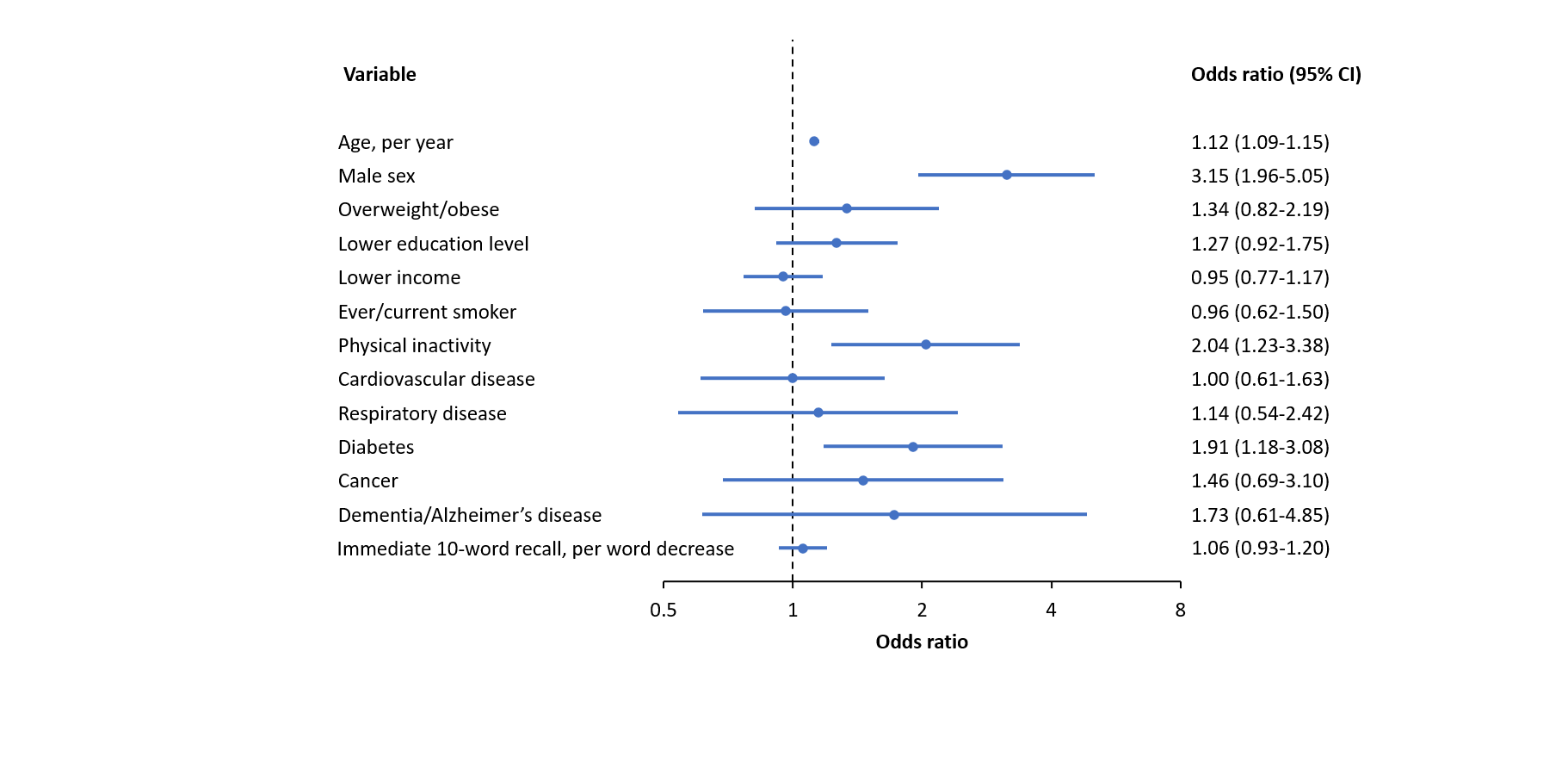

SHARE, the Survey of Health, Ageing and Retirement in Europe.

**Table S1. Characteristics of the study sample and excluded persons in SHARE**

| **Characteristic** | **Study sample**  **(n=4,062)** | **Excluded persons**  **(n=72,790)^#^** | ***P*** |
| --- | --- | --- | --- |
| Age, median (IQR), y | 67.0 (62.0-74.0) | 71.0 (64.0-79.0) | <0.0001 |
| Age category, y |  |  |  |
| 50-60 | 640 (15.8%) | 7,830 (10.8%) | <0.0001 |
| 60-70 | 1,773 (43.7%) | 24,477 (33.6%) |  |
| 70-80 | 1,139 (28.0%) | 23,511 (32.3%) |  |
| ≥80 | 510 (12.6%) | 16,965 (23.3%) |  |
| Sex |  |  |  |
| Male | 1,660 (40.9%) | 31,414 (43.2%) | 0.004 |
| Female | 2,402 (59.1%) | 41,376 (56.8%) |  |
| Race |  |  |  |
| White | NA | NA | NA |
| Other | NA | NA |  |
| Body mass index, kg/m^2^ |  |  |  |
| Not overweight or obese: <25 | 1,225 (30.2%) | 24,310 (34.9%) | <0.0001 |
| Overweight or obese: ≥25 | 2,837 (69.8%) | 45,430 (65.1%) |  |
| Educational level |  |  |  |
| Low | 1,231 (30.3%) | 26,688 (36.9%) | <0.0001 |
| Medium | 1,841 (45.3%) | 30,219 (41.7%) |  |
| High | 990 (24.4%) | 15,517 (21.4%) |  |
| Household income |  |  |  |
| Quartile 1 (lowest) | 795 (19.6%) | 19,273 (26.5%) | <0.0001 |
| Quartile 2 | 991 (24.4%) | 18,854 (25.9%) |  |
| Quartile 3 | 1,074 (26.4%) | 18,021 (24.8%) |  |
| Quartile 4 (highest) | 1,202 (29.6%) | 16,642 (22.9%) |  |
| Ever or current smoker | 1,708 (42.1%) | 27,151 (43.1%) | 0.180 |
| Physical inactivity | 374 (9.2%) | 8,752 (13.9%) | <0.0001 |
| Cardiovascular disease | 548 (13.5%) | 11,288 (15.6%) | <0.0001 |
| Respiratory disease | 213 (5.2%) | 3,967 (5.5%) | 0.502 |
| Diabetes | 510 (12.6%) | 10,099 (14.0%) | 0.011 |
| Cancer | 175 (4.3%) | 3,593 (5.0%) | 0.057 |
| Dementia or Alzheimer’s disease | 42 (1.0%) | 1,841 (2.6%) | <0.0001 |

Values are median (IQR) or n (%). Characteristics among the study sample and excluded persons were tested using the Mann-Whitney test or the chi-square test. SHARE, the Survey of Health, Ageing and Retirement in Europe.

^#^ Persons were excluded if they did not have 10-words recall tests at baseline, did not have COVID-19 infection, were younger than 50 years in 2020, or had missing values for covariates (including age, sex, body mass index, smoking status, physical activity, household income, education, and underlying health conditions such as cardiovascular disease, respiratory disease, diabetes, cancer, and dementia or Alzheimer’s disease).

**Table S2. Characteristics of the study sample and excluded persons in HRS**

| **Characteristic** | **Study sample**  **(n=1,349)** | **Excluded persons**  **(n=15,494) ^#^** | ***P*** |
| --- | --- | --- | --- |
| Age, median (IQR), y | 64.0 (59.0-71.0) | 68.0 (60.0-79.0) | <0.0001 |
| Age category, y |  |  |  |
| 50-60 | 392 (29.1%) | 3,368 (21.7%) | <0.0001 |
| 60-70 | 552 (40.9%) | 5,108 (33.0%) |  |
| 70-80 | 259 (19.2%) | 3,479 (22.5%) |  |
| ≥80 | 146 (10.8%) | 3,538 (22.8%) |  |
| Sex |  |  |  |
| Male | 566 (42.0%) | 6,390 (41.2%) | 0.610 |
| Female | 783 (58.0%) | 9,103 (58.8%) |  |
| Race |  |  |  |
| White | 903 (66.9%) | 10,235 (66.1%) | 0.512 |
| Other | 446 (33.1%) | 5,259 (33.9%) |  |
| Body mass index, kg/m^2^ |  |  |  |
| Not overweight or obese: <25 | 261 (19.4%) | 4,103 (26.9%) | <0.0001 |
| Overweight or obese: ≥25 | 1,088 (80.7%) | 11,158 (73.1%) |  |
| Educational level |  |  |  |
| Low | 217 (16.1%) | 2,953 (19.1%) | 0.019 |
| Medium | 417 (30.9%) | 4,493 (29.1%) |  |
| High | 715 (53.0%) | 7,979 (51.7%) |  |
| Household income |  |  |  |
| Quartile 1 (lowest) | 282 (20.9%) | 3,946 (25.5%) | <0.0001 |
| Quartile 2 | 289 (21.4%) | 3,954 (25.5%) |  |
| Quartile 3 | 374 (27.7%) | 3,814 (24.6%) |  |
| Quartile 4 (highest) | 404 (30.0%) | 3,780 (24.4%) |  |
| Ever or current smoker | 691 (51.2%) | 8,353 (54.1%) | 0.040 |
| Physical inactivity | 203 (15.1%) | 3,315 (21.4%) | <0.0001 |
| Cardiovascular disease | 335 (24.8%) | 4,650 (30.0%) | <0.0001 |
| Respiratory disease | 135 (10.0%) | 1,813 (11.7%) | 0.062 |
| Diabetes | 388 (28.8%) | 4,536 (29.3%) | 0.691 |
| Cancer | 187 (13.9%) | 2,450 (15.8%) | 0.059 |
| Dementia or Alzheimer’s disease | 24 (1.8%) | 712 (4.6%) | <0.0001 |

Values are median (IQR) or n (%). Characteristics among the study sample and excluded persons were tested using the Mann-Whitney test or the chi-square test. HRS, the Health and Retirement Study.

^#^ Persons were excluded if they did not have 10-words recall tests at baseline, did not have COVID-19 infection, were younger than 50 years in 2020, or had missing values for covariates (including age, sex, race, body mass index, smoking status, physical activity, household income, education, and underlying health conditions such as cardiovascular disease, respiratory disease, diabetes, cancer, and dementia or Alzheimer’s disease).

**Table S3. Characteristics of the study sample and excluded persons in ELSA**

| **Characteristic** | **Study sample**  **(n=278)** | **Excluded persons**  **(n=8312) ^#^** | ***P*** |
| --- | --- | --- | --- |
| Age, median (IQR), y | 66.0 (57.0-73.0) | 69.0 (62.0-77.0) | <0.0001 |
| Age category, y |  |  |  |
| 50-60 | 88 (31.7%) | 1688 (20.3%) | <0.0001 |
| 60-70 | 100 (36.0%) | 2537 (30.5%) |  |
| 70-80 | 58 (20.9%) | 2572 (30.9%) |  |
| ≥80 | 32 (11.5%) | 1515 (18.2%) |  |
| Sex |  |  |  |
| Male | 102 (36.7%) | 3715 (44.7%) | 0.008 |
| Female | 176 (63.3%) | 4597 (55.3%) |  |
| Race |  |  |  |
| White | 265 (95.3%) | 7909 (95.2%) | 0.512 |
| Other | 13 (4.7%) | 397 (4.8%) |  |
| Body mass index, kg/m^2^ |  |  |  |
| Not overweight or obese: <25 | 85 (30.6%) | 2305 (30.5%) | 0.970 |
| Overweight or obese: ≥25 | 193 (69.4%) | 5260 (69.5%) |  |
| Educational level |  |  |  |
| Low | 55 (20.0%) | 2080 (27.6%) | 0.005 |
| Medium | 64 (23.0%) | 1822 (24.2%) |  |
| High | 159 (57.2%) | 3623 (48.2%) |  |
| Household income |  |  |  |
| Quartile 1 (lowest) | 60 (21.6%) | 2071 (25.4%) | <0.0001 |
| Quartile 2 | 56 (20.1%) | 2062 (25.3%) |  |
| Quartile 3 | 62 (22.3%) | 2045 (25.1%) |  |
| Quartile 4 (highest) | 100 (36.0%) | 1975 (24.2%) |  |
| Ever or current smoker | 159 (57.2%) | 4918 (59.2%) | 0.510 |
| Physical inactivity | 49 (17.6%) | 1593 (19.2%) | 0.520 |
| Cardiovascular disease | 22 (7.9%) | 840 (10.1%) | 0.231 |
| Respiratory disease | 19 (6.8%) | 572 (6.9%) | 0.976 |
| Diabetes | 34 (12.2%) | 1102 (13.3%) | 0.618 |
| Cancer | 34 (12.2%) | 1110 (13.4%) | 0.587 |
| Dementia or Alzheimer’s disease | 1 (0.4%) | 208 (2.5%) | 0.023 |

Values are median (IQR) or n (%). Characteristics among the study sample and excluded persons were tested using the Mann-Whitney test or the chi-square test. ELSA, the English Longitudinal Study of Ageing.

^#^ Persons were excluded if they did not have 10-words recall tests at baseline, did not have COVID-19 infection, were younger than 50 years in 2020, or had missing values for covariates (including age, sex, race, body mass index, smoking status, physical activity, household income, education, and underlying health conditions such as cardiovascular disease, respiratory disease, diabetes, cancer, and dementia or Alzheimer’s disease).

**Table S4. Sensitivity analysis of associations of 10-words recall with COVID-19 hospitalization in SHARE and HRS after excluding participants whose diagnosis of COVID-19 infection was based on symptoms alone**

| **Model** | **SHARE cohort, N=2823^*^** | | **HRS cohort, N=1176^#^** | |
| --- | --- | --- | --- | --- |
|  | **OR (95% CI)** | ***P*** | **OR (95% CI)** | ***P*** |
| Immediate 10-words recall | | | | |
| Model 0 | 1.28 (1.22-1.35) | <0.001 | 1.24 (1.11-1.39) | <0.001 |
| Model 1 | 1.16 (1.10-1.23) | <0.001 | 1.15 (1.02-1.29) | 0.018 |
| Model 2 | 1.15 (1.09-1.22) | <0.001 | 1.10 (0.97-1.24) | 0.136 |
| Model 3 | 1.15 (1.08-1.22) | <0.001 | 1.08 (0.95-1.22) | 0.239 |
| Delayed 10-words recall | | | | |
| Model 0 | 1.23 (1.17-1.28) | <0.001 | 1.25 (1.14-1.37) | <0.001 |
| Model 1 | 1.12 (1.07-1.18) | <0.001 | 1.18 (1.07-1.30) | 0.001 |
| Model 2 | 1.12 (1.06-1.17) | <0.001 | 1.15 (1.03-1.27) | 0.010 |
| Model 3 | 1.11 (1.05-1.17) | <0.001 | 1.12 (1.01-1.25) | 0.029 |

^*^ Of the 2823 participants with COVID-19 infection, 610 were hospitalized in the SHARE cohort.

^#^ Of the 1176 participants with COVID-19 infection, 142 were hospitalized in the HRS cohort.

Model 0: unadjusted.

Model 1: adjusted for age, sex, and race.

Model 2: further adjusted for smoking status, physical activity, body mass index, educational level, and household income.

Model 3: further adjusted for cardiovascular disease, respiratory disease, diabetes, cancer, and dementia or Alzheimer’s disease.

Race was not included in SHARE. Dementia or Alzheimer's disease was not adjusted in ELSA as only one participant had the disease.

HRS, the Health and Retirement Study; SHARE, the Survey of Health, Ageing and Retirement in Europe.

**Table S5.** **Sensitivity analysis of associations of 10-words recall with COVID-19 mortality in the SHARE cohort after excluding participants whose diagnosis of COVID-19 infection was based on symptoms only (N=2823)^*^**

| **Model** | **OR (95% CI)** | ***P*** |
| --- | --- | --- |
| Immediate 10-words recall | | |
| Model 0 | 1.36 (1.22-1.51) | <0.001 |
| Model 1 | 1.10 (0.98-1.24) | 0.114 |
| Model 2 | 1.07 (0.94-1.21) | 0.335 |
| Model 3 | 1.05 (0.92-1.20) | 0.441 |
| Delayed 10-words recall | | |
| Model 0 | 1.39 (1.25-1.54) | <0.001 |
| Model 1 | 1.18 (1.05-1.32) | 0.005 |
| Model 2 | 1.15 (1.02-1.30) | 0.018 |
| Model 3 | 1.15 (1.01-1.29) | 0.029 |

^*^ Of the 2823 participants with COVID-19 infection in the SHARE cohort, 102 died of COVID-19 or related complications.

Model 0: unadjusted.

Model 1: adjusted for age and sex.

Model 2: further adjusted for smoking status, physical activity, body mass index, educational level, and household income.

Model 3: further adjusted for cardiovascular disease, respiratory disease, diabetes, cancer, and dementia or Alzheimer's disease.

SHARE, the Survey of Health, Ageing and Retirement in Europe.

**Table S6. Sensitivity analysis of associations of 10-words recall with COVID-19 hospitalization in SHARE and HRS** **after excluding participants with dementia or Alzheimer's disease**

| **Model** | **SHARE cohort, N=4020** | | **HRS cohort, N=1325** | |
| --- | --- | --- | --- | --- |
|  | **OR (95% CI)** | ***P*** | **OR (95% CI)** | ***P*** |
| Immediate 10-words recall | | | | |
| Model 0 | 1.30 (1.24-1.37) | <0.001 | 1.22 (1.09-1.36) | 0.001 |
| Model 1 | 1.18 (1.12-1.25) | <0.001 | 1.13 (1.00-1.27) | 0.051 |
| Model 2 | 1.16 (1.09-1.23) | <0.001 | 1.07 (0.94-1.21) | 0.311 |
| Model 3 | 1.15 (1.09-1.22) | <0.001 | 1.07 (0.94-1.21) | 0.305 |
| Delayed 10-words recall | | | | |
| Model 0 | 1.23 (1.18-1.30) | <0.001 | 1.25 (1.13-1.37) | <0.001 |
| Model 1 | 1.14 (1.09-1.19) | <0.001 | 1.18 (1.06-1.30) | 0.002 |
| Model 2 | 1.12 (1.06-1.17) | <0.001 | 1.14 (1.02-1.26) | 0.019 |
| Model 3 | 1.11 (1.06-1.17) | <0.001 | 1.14 (1.02-1.27) | 0.018 |

Model 0: unadjusted.

Model 1: adjusted for age, sex, and race.

Model 2: further adjusted for smoking status, physical activity, body mass index, educational level, and household income.

Model 3: further adjusted for cardiovascular disease, respiratory disease, diabetes, and cancer.

Race was not included in SHARE.

HRS, the Health and Retirement Study; SHARE, the Survey of Health, Ageing and Retirement in Europe.

**Table S7. Sensitivity analysis of associations of 10-words recall with COVID-19 mortality in SHARE after excluding participants with dementia or Alzheimer's disease (n=4020)**

| **Model** | **OR (95% CI)** | ***P*** |
| --- | --- | --- |
| Immediate 10-words recall | | |
| Model 0 | 1.38 (1.24-1.54) | <0.001 |
| Model 1 | 1.12 (0.99-1.27) | 0.078 |
| Model 2 | 1.07 (0.94-1.22) | 0.302 |
| Model 3 | 1.07 (0.94-1.22) | 0.332 |
| Delayed 10-words recall | | |
| Model 0 | 1.40 (1.26-1.55) | <0.001 |
| Model 1 | 1.19 (1.06-1.33) | 0.003 |
| Model 2 | 1.16 (1.03-1.30) | 0.017 |
| Model 3 | 1.15 (1.02-1.30) | 0.020 |

Model 0: unadjusted.

Model 1: adjusted for age and sex.

Model 2: further adjusted for smoking status, physical activity, body mass index, educational level, and household income.

Model 3: further adjusted for cardiovascular disease, respiratory disease, diabetes, and cancer.

SHARE, the Survey of Health, Ageing and Retirement in Europe.

**Table S8. Sensitivity analyses after imputation for missing covariates in SHARE and HRS**

|  | SHARE cohort, N=4562 | | HRS cohort, N=1366 | |
| --- | --- | --- | --- | --- |
|  | OR (95% CI) | *P* | OR (95% CI) | *P* |
| Immediate 10-words recall |  |  |  |  |
| Hospitalization | 1.15 (1.09-1.21) | <0.001 | 1.06 (0.94-1.20) | 0.348 |
| Mortality | 1.01 (0.90-1.15) | 0.817 | NA | NA |
| Delayed 10-words recall |  |  |  |  |
| Hospitalization | 1.11 (1.06-1.17) | <0.001 | 1.11 (1.00-1.23) | 0.045 |
| Mortality | 1.09 (0.98-1.22) | 0.121 | NA | NA |

Adjusted for age, sex, race (HRS only), smoking status, physical activity, body mass index, educational level, household income, cardiovascular disease, respiratory disease, diabetes, cancer, and dementia or Alzheimer's disease.

HRS, the Health and Retirement Study; SHARE, the Survey of Health, Ageing and Retirement in Europe.
